# Higher Brain Grey Matter Density in Mild to Moderate Chronic Low Back Pain Patients

**DOI:** 10.1101/2025.01.07.25320128

**Authors:** Monica Sean, Samantha Cote, Alexia-Coulombe-Lévêque, Julia Huck, Marylie Martel, Guillaume Léonard, Kevin Whittingstall, Pascal Tétreault

## Abstract

Chronic low back pain (CLBP) is the leading cause of disability worldwide. Structural abnormalities in the lumbar region rarely account for the pain symptoms observed. In contrast, the brain plays a central role in chronic pain, with several studies suggesting that chronic pain may result from the persistence of pain memories and/or the inability to extinguish these memories following an initial inciting injury. Previous research has reported a reduction in grey matter density (GMD) in CLBP patients, as measured by magnetic resonance imaging (MRI). Notably, lower GMD has been observed in patients with moderate to severe CLBP who are undergoing prescribed pharmacological treatments. However, it remains unclear whether these differences in GMD could be associated with a less severe condition.

This study aimed to investigate whether GMD is altered in a cohort with mild to moderate CLBP symptoms, and who have not been prescribed pharmacological treatments. To achieve this, we acquired T1-weighted MRI scans from 25 healthy controls (HC) and 27 untreated individuals with mild to moderate CLBP. Scans were taken at baseline, 2 months, and 4 months after baseline. GMD analysis was carried out using the FMRIB Software.

Our results consistently showed higher GMD in the CLBP group compared to HC. These findings suggest that the observed alterations in GMD may be related to the condition itself, and can occur even in patients with milder symptoms and better physical function, without the influence of opioids, anticonvulsants, or antidepressants. Given that our participants differ from those typically studied in the literature, our results imply that they may be in a distinct brain state relative to the condition. Further research is needed to elucidate the underlying biological mechanisms and confirm whether these brain alterations are indeed a characteristic feature of CLBP.

## 1. Introduction

Approximately one-fifth of Canadians experience chronic pain (CP) (Canadian Pain Task Force, 2019); across all CP conditions, chronic low back pain (CLBP) represents one of the leading causes of disability worldwide (Vos et al., 2017). In 90% of CLBP cases, the pain cannot be linked to a specific musculoskeletal cause (Finucane et al., 2020; Kahere & Ginindza, 2020; Kasch et al., 2022a; Maher et al., 2017; Nijs et al., 2024). Several studies have found no association between imaging abnormalities of the dorsal-lumbar regions and the presence of pain (Brinjikji, Diehn, et al., 2015; Brinjikji, Luetmer, et al., 2015; Burgstaller et al., 2016; Hall et al., 2021; Kasch et al., 2022b). This is likely due to the fact that several morphological abnormalities in the spine can be present in individuals with and without pain; representing normal anatomical variations or nonpathological degenerative changes associated with aging that begin in early adulthood (Benoist, 2003; Brinjikji, Luetmer, et al., 2015; Parenteau et al., 2021; Wocial et al., 2021).

The brain plays a central role in the perception and processing of pain, particularly in the development of CP (Apkarian, 2008; Bushnell et al., 2013; Garland, 2012). Several researchers suggest that CP may result from the persistence of pain memories and/or the inability to extinguish the memory of pain triggered by an initial injury (Apkarian, 2008; Mansour et al., 2014; McCarberg & Peppin, 2019; Phelps et al., 2021). Indeed, numerous studies using magnetic resonance imaging (MRI) have reported structural brain abnormalities in individuals with CLBP (Apkarian et al., 2009; Baliki et al., 2011; Biggs et al., 2022; Makary et al., 2020; Mansour et al., 2014; Phelps et al., 2021; Simons et al., 2014; Vachon-Presseau, 2018; Yi & Zhang, 2011). Most studies on grey matter density (GMD) have reported lower GMD in patients with CLBP (Apkarian et al., 2004; Baliki et al., 2011; Ivo et al., 2013; Schmidt-Wilcke et al., 2006; Ung et al., 2014; Yu et al., 2021). These studies, however, primarily focused on individuals with moderate to severe pain or those who used prescribed pharmacological treatments to manage their symptoms. Since these treatments are known to influence cerebral grey matter volume (Puiu et al., 2016; Russell et al., 2018; Younger et al., 2011), it is challenging to determine whether the observed reductions in GMD are due to the CP condition itself or the effects of these centrally acting pharmacological agents. For example, Younger et al. (2011) demonstrated long-lasting reduced grey matter volume in several brain regions after just one month of morphine treatment (Younger et al., 2011). Moreover, these studies typically involved patients with more severe pain symptoms, raising the question of whether similar changes in grey matter occur in individuals with milder symptoms.

We conducted an observational study to assess pain symptoms at three time points over a 4-month period in individuals with CLBP who had never used opioids, anticonvulsants, or antidepressants, as well as in HC. The results from pain questionnaires, which were analyzed and discussed in a previous publication (Sean et al., 2024), revealed that our study participants experienced mild to moderate pain symptoms. This presented an opportunity to investigate grey matter density (GMD) in a cohort with milder symptoms and better physical function than those typically studied in the literature.

In the present study, we evaluated GMD using high-resolution T1-weighted MRI scans across three visits over 4 months in both the mild CLBP cohort and HC. The primary aim of this study was to identify regions where GMD might differ between individuals with CLBP who had not used opioids, anticonvulsants, or antidepressants, substances known to influence grey matter structure.

## 2. Methods

### 2.1 Participants

All participants provided written informed consent for their participation into the study. Ethics approval was granted from the institutional review board of the Centre intégré universitaire de santé et de services sociaux de l’Estrie - Centre hospitalier universitaire de Sherbrooke (CIUSSS de l’Estrie-CHUS) (Sherbrooke, Quebec, Canada; approval #2021-3861). The study has been registered on Open Science Framework (OSF), under the name “Pilot project on brain and lower back imaging of chronic pain” (https://osf.io/p2z6y) and was conducted in accordance with the declaration of Helsinki.

Participants were recruited using posters at the CIUSSS de l’Estrie-CHUS, local physiotherapy clinics, Facebook ads, and by word of mouth. Twenty-seven CLBP patients and 25 HC aged 18 to 75 years old took part in this study. HC were matched with CLBP patients for sex and age. Detailed inclusion/exclusion criteria are discussed elsewhere (https://osf.io/p2z6y<u>) (Sean et al., 2024)</u>, but briefly, inclusion criteria for the CLBP were: 1) low back pain (≥ 4 months) with or without pain radiating to the legs or radiating to the neck; 2) average pain intensity of ≥ 3/10 in the 24-hour period before the initial visit; 3) pain primarily localized in the lower back; 4) no history/no use of opioids, anticonvulsants or antidepressants. Exclusion criteria for HC were: 1) history of chronic pain; 2) pain at the time of testing; 3) an outstanding painful episode within 3 months of enrollment in the study. Exclusion criteria for the two populations included: 1) neurological, cardiovascular, or pulmonary disorders; 2) comorbid pain syndrome (i.e., fibromyalgia, osteoarthritis, irritable bowel syndrome, migraine etc.); 3) history of surgical intervention in the back; 4) a corticosteroid infiltration within the past year; 6) pregnancy (current or planned during the course of the study); 7) inability to read or understand French; 8) contra-indication to MRI.

### 2.2 Study design

The study had an observational longitudinal design. All participants attended three sessions at the Centre de recherche du CHUS: baseline (BL), 2-months (2M) and 4-months (4M) after BL. During each visit, participants completed an array of pain questionnaires and underwent brain MRI. Participants were scanned three times over a four-month period without any treatment or intervention.

### 2.3 MRI acquisition

MRI data was collected on a 3 Tesla (3T) Ingenia scanner equipped with a 32- channel head-coil (Philips Healthcare, Best, Netherlands). A high-resolution 3D gradient-echo T1 weighted image was first acquired (flip angle of 8°, TR/TE of 7.9/3.5 ms, 1 mm isotropic voxels with a field of view (FOV) of 224 × 224 × 160 mm). The total scan time was 4 minutes.

### 2.4 Preprocessing of T1 weighted images

T1-weighted images were converted from DICOM to NIFTI using the dcm2niix tool embedded into MRIcroGL software (https://www.nitrc.org/projects/mricrogl/). Voxel Based Morphometry (VBM) analysis was carried out using the default parameters within the FMRIB Software Library (FSL-VBM) pipeline (Douaud et al., 2007). A detailed description is presented in **Figure 1**. Briefly, the preprocessing steps involved skull stripping and segmentation of the grey matter (GM), white matter (WM) and cerebrospinal fluid (CSF). Next, a study-specific GM template was created by combing HC and CLPB participants. First, GM images were aligned using an affine registration to the standard GM template based on the ICBM-152 stereotaxic atlas (FSL version 6.0.6.1 (Andersson et al., 2007)), creating a GM template. Second, GM images are registered to the GM template using non-linear registration, creating another template. Third, both templates are averaged to create a final study-specific template. After, all GM maps of our participants are non-linearly registered to this final study-specific template. Each voxel of each registered GM image is multiplied by the Jacobian of the warp field and then smoothed by an isotropic Gaussian kernel with a sigma value of 2 mm (S2). Because smoothing can influence statistical results from VBM approaches, we compared results at an isotropic Gaussian kernel with a sigma value of 3 mm (S3) and 4 mm (S4) (Coalson et al., 2018).

**Figure 1:**
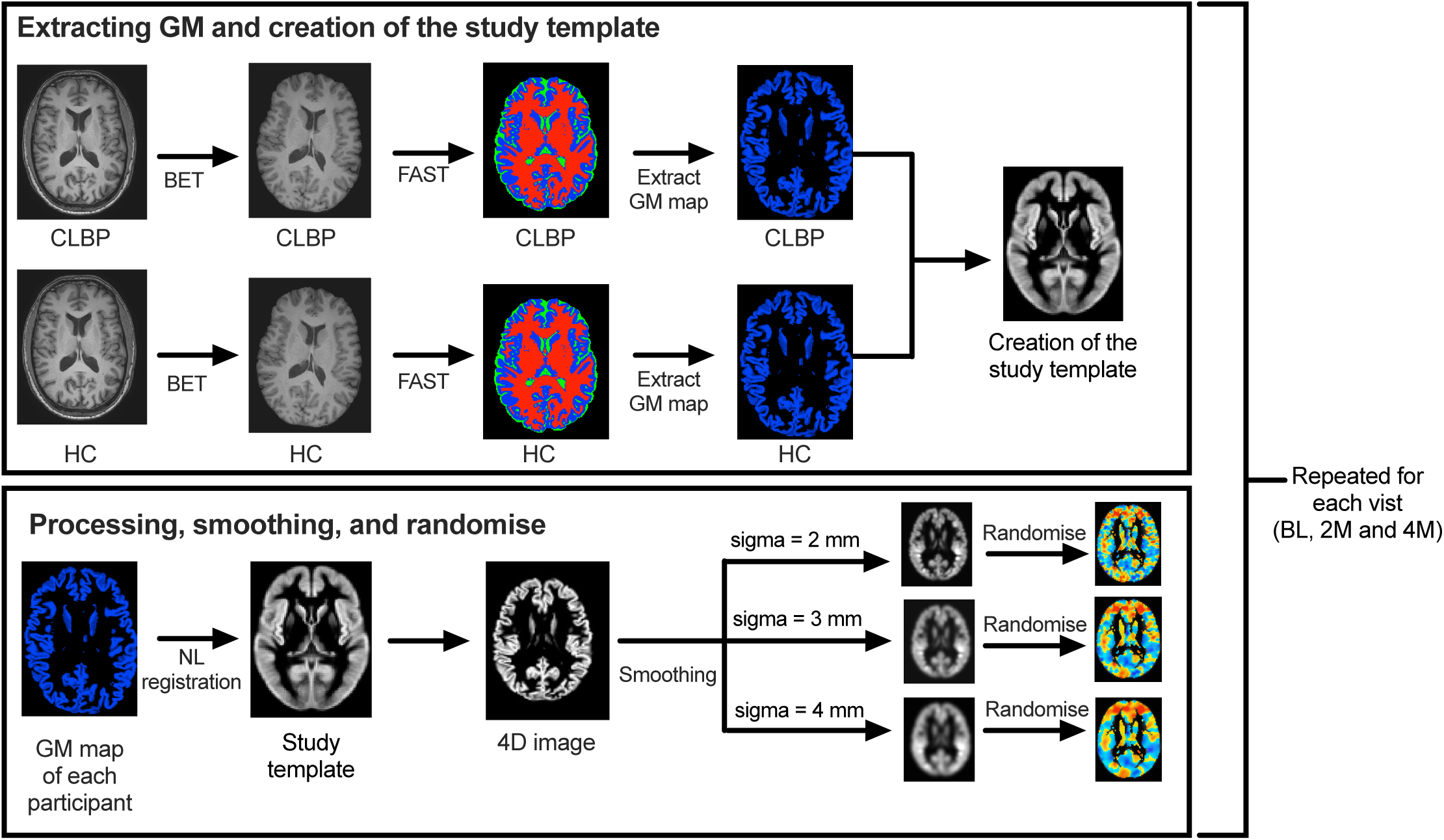
Overview of the FSL-VBM pipeline CLBP = chronic low back pain, HC = healthy controls, BL= baseline, 2M = 2-months after baseline, 4M = 4 months after baseline, GM = grey matter, BET = Brain Extraction Tool, FAST = FMRIB’s Automated Segmentation Tool, NL = Non-Linear registration

### 2.5 VBM and statistical analysis

FSL *Randomize* (Winkler et al., 2014) was used to conduct a voxel-wise general linear model (GLM) to examine the differences between groups. Non-parametric testing with 5000 random permutations were performed on the whole-brain GM maps to create t-test maps between HC and CLBP.

To isolate brain regions with significant differences between HC and CLBP patients, four different thresholds were applied using AFNI (Cox, 1996) (**Figure 2**).

**Figure 2:**
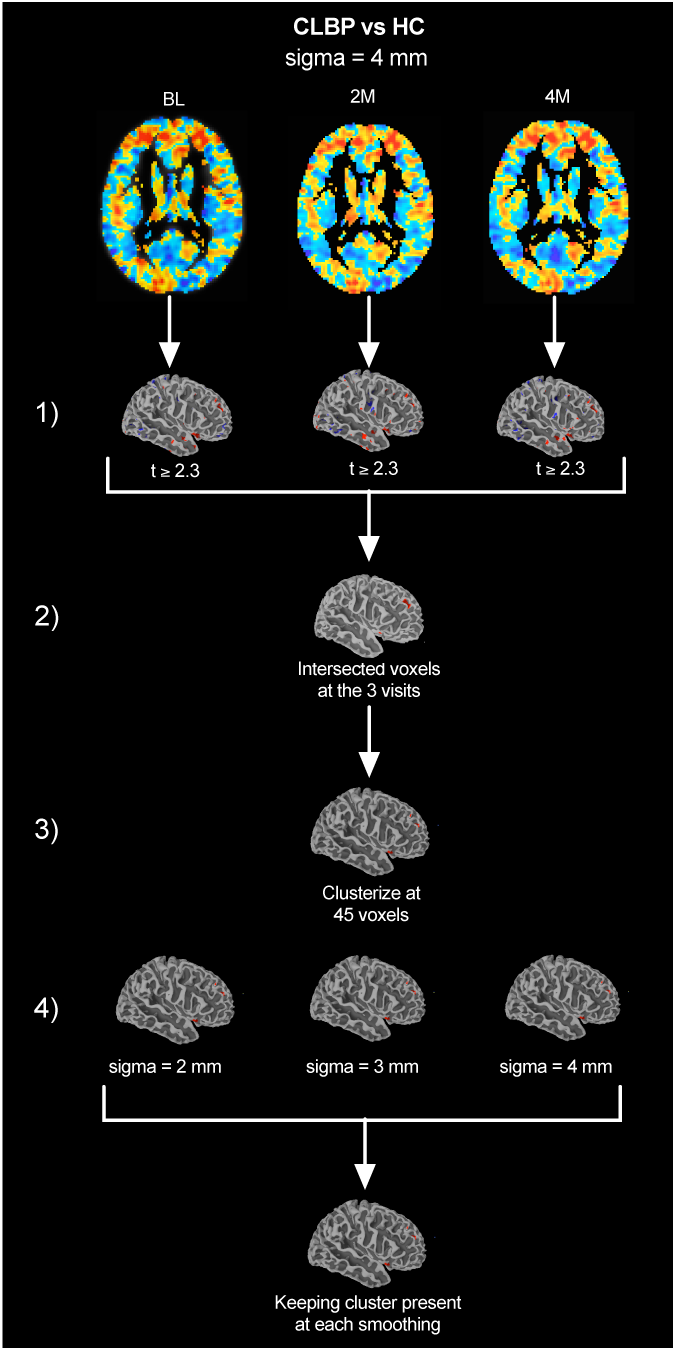
Overview of the four different thresholding analyses. Thresholding’s are based on 1) t-stats maps obtained from randomize; 2) significant and overlapping voxels present at each visit; 3) clusters of at least 45 voxels and 4) clusters that were present in each smoothing kernels. BL= baseline, 2M = 2-months after baseline, 4M = 4 months after baseline.

First, in each visit (BL, 2M and 4M), only voxels with independent t-test score higher or greater than 2.3 were kept from the randomized t-test maps. Second, across these remaining voxels, we created an intersection map of only voxels present at each of the three visits (BL, 2M and 4M) with independent t-test score higher or greater than 2.3 greater were retained. Third, this intersection map was subject to 3 different cluster sizes: 15 voxels, 30 voxels and 45 voxels, wherein voxels face must touch (first-nearest neighbor clustering), meaning that clusters present at 45 voxels were retained. Finally, clusters present at each smoothing (sigma=2, sigma=3, and sigma=4) were retained based on the approximate localization (z, y, and z MNI coordinates) of the cluster using Brainnetome Atlas (https://atlas.brainnetome.org/bnatlas.html). Clusters passing the four different thresholding were considered as the most significant clusters. Average GMD values of most significant clusters were extracted at each visit (probability values ranging from 0 to 1, wherein 0 = no GM is present and 1 = filled with GM).

Intracranial volume (ICV) was obtained using FreeSurfer (version 7.2 (http://surfer.nmr.mgh.harvard.edu/). ICV was compared between both groups using a Student’s t-test (unpaired).

Sociodemographic data were also compared using a Student’s t-test (unpaired) or Chi-square test with SPSS (version 28).

### 2.6 Pain questionnaires

As mentioned before, pain and other behavioral questionnaires have been analyzed and discussed in a previous article (Sean et al., 2024). Briefly, CLBP patients completed eight pain and behavioral questionnaires online using the platform “Research Electronic Data Capture” (REDCap). Data from the Brief Pain Inventory (BPI) short form subscales (BPI severity and BPI interference) are presented in **Table 1**. The measure “average intensity of pain in the last 24 hours” used here was also computed from the BPI severity score.

**Table 1:** Characteristics of the sample.

|  | CLBP | HC | p-value |
| --- | --- | --- | --- |
| BL | 27 | 25 | - |
| 2M | 24 | 25 | - |
| 4M | 23 | 25 | - |
| Intracranial volume (ICV) (cm<sup>3</sup>) |  |  |  |
| BL | 1530.31 ± 155.30 | 1532.76 ± 154.37 | p=0.955 |
| 2M | 1537.03 ± 163.59 | 1530.78 ± 152.40 | p=0.890 |
| 4M | 1541.83 ± 153.97 | 1535.64 ± 153.77 | p=0.890 |
| Women | 13 | 15 | p=0.419 |
| Men | 14 | 10 |  |
| Age (average ± std) | 43 ± 16 | 42 ± 14 | p=0.700 |
| Ethnicity |  |  |  |
| Caucasian | 20 | 24 | p=0.178 |
| Asian | 1 | 1 |  |
| Hispanic | 4 | - |  |
| African | 1 | - |  |
| Arabic | 1 | - |  |
| Education level |  |  |  |
| Primary school | - | - | p=0.318 |
| High school | - | - |  |
| Apprenticeship | 5 | 2 |  |
| College | 8 | 6 |  |
| University | 14 | 17 |  |
| Annual income |  |  |  |
| less than 20K | 3 | 5 |  |
| 20K - 35K | 6 | 2 |  |
| 35K - 50K | 7 | 1 | p=0.186 |
| 50K - 65K | 5 | 6 |  |
| 65K - 80K | 2 | 5 |  |
| 80K - 100K | 3 | 4 |  |
| 100K and more | 1 | 2 |  |
| **Average intensity of pain in the last 24 hours (0 – 10)** |  |  | NA |
| **BL** | 4 ± 1 | NA |  |
| **2M** | 4 ± 1 | NA |  |
| **4M** | 4 ± 4 | NA |  |
| **Brief Pain Inventory - severity (0 – 40)** |  |  | NA |
| **BL** | 17 ± 4 | NA |  |
| **2M** | 18 ± 6 | NA |  |
| **4M** | 15 ± 7 | NA |  |
| **Brief Pain Inventory - interference (0 – 70)** |  |  | NA |
| **BL** | 17 ± 10 | NA |  |
| **2M** | 15 ± 11 | NA |  |
| **4M** | 9 ± 7 | NA |  |
| **Pain duration** |  |  | NA |
| 4 month - 5 months | 0 | NA |  |
| 6 months - 12 month | 5 | NA |  |
| 1 - 4 years | 8 | NA |  |
| 5 years and more | 14 | NA |  |

## 3. Results

### 3.1 Participants

Fifty-two participants (25 HC and 27 CLBP) were recruited in the study. Three CLBP participants dropped out after the first visit (unexpected pregnancy [n=1], discomfort during MRI [n=1], scheduling conflicts [n=1]), and one dropped out after the second visit (move to a different city [n=1]). All HC completed the three experimental visits. The 25 HC (15 women, 10 men) were aged 42 ± 14 years old, and the 27 CLBP participants (13 women, 14 men) were aged 43 ± 16 years old. All variables showed no significant difference between both groups.

Results from the pain questionnaires, for example from the last pain intensity in the last 24 hours, BPIs and BPIi, showed that our sample consisted of patients with mild to moderate pain symptoms (Cleeland & Ryan, 1994; Dworkin et al., 2012; Poquet & Lin, 2016). Questionnaire scores and characteristics of the sample are presented in **Table 1**.

### 3.2 CLBP patients show higher GMD

Initial VBM results showed differences in GMD between groups at each visit (BL, 2M and 4M). Across all the statistically significant voxels, 76%, 81% and 71% of the voxels showed higher GMD in CLBP compared to HC, respectively at BL, 2M and 4M when using a smoothing kernel of 4 mm as shown in **Figure 3**. Similar results were obtained when using different smoothing kernels (sigma = 2 mm and sigma = 3 mm; see **Supplementary 1 and Supplementary 2)**.

**Figure 3:**
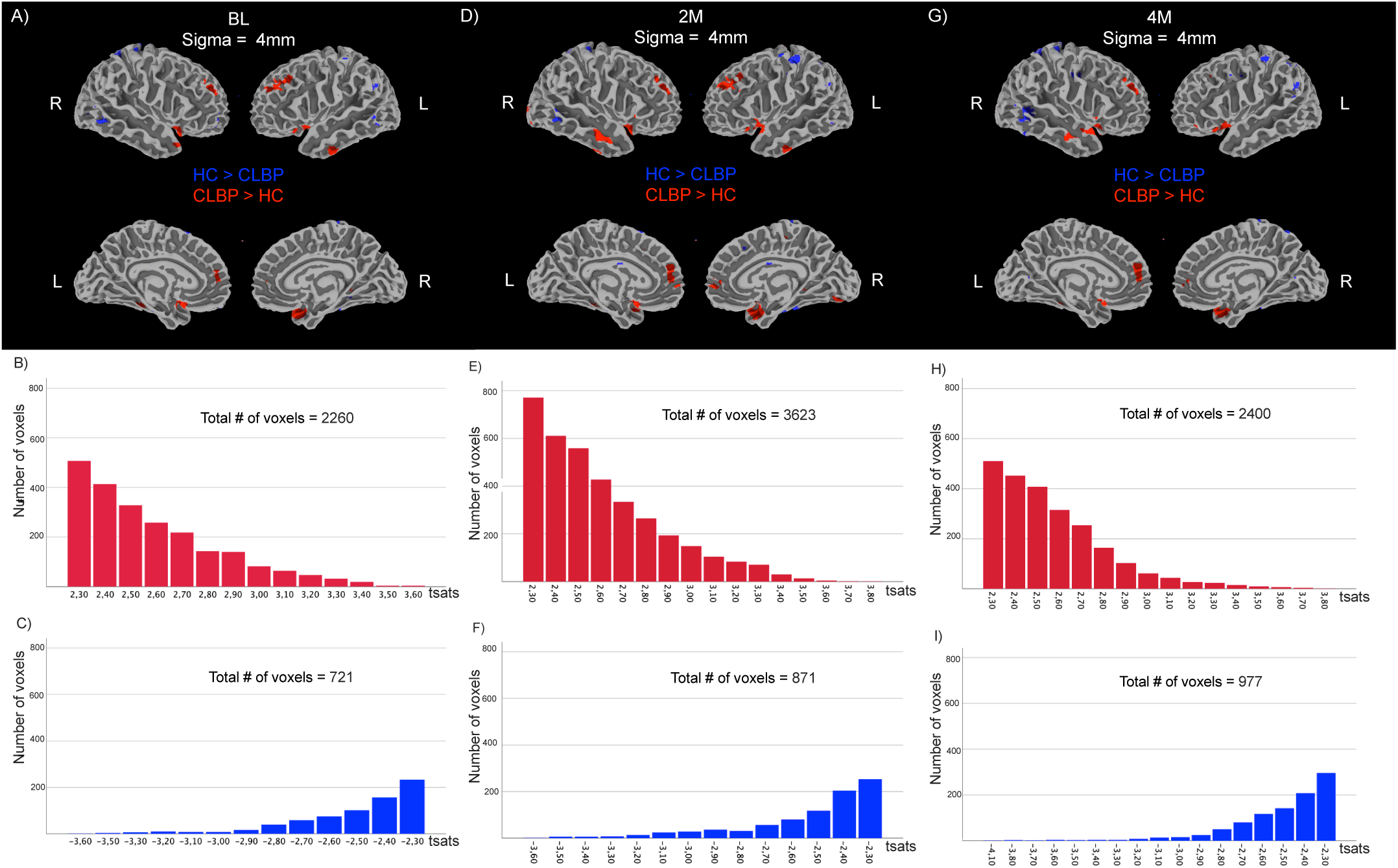
CLBP patients show higher GMD GMD differences (sigma = 4mm) between groups are shown at A) BL, B) 2M and C) 4M. Voxels where GMD is lower in the CLBP group compared to HC are shown in blue. Voxels where GMD is higher in the CLBP group compared to HC are shown in red. For each map, two bar charts are presented to show the number of significant voxels for each t-statistics. The red bar chart (B, E, and H) indicates the number of voxels where GMD is higher in the CLBP group compared to HC (t ≥ 2.3) at BL, 2M, and 4 M respectively. The blue chart (C, F, and I) indicated the number of voxels where GMD is lower in the CLBP group compared to HC (t ≥ - 2.3) at BL, 2M, and 4 M respectively.

### 3.3 Higher GMD was specifically observed in the frontal and medial temporal lobe

After keeping only voxels that intersected at all three visits (BL, 2M, and 4M), the analysis identified 16 clusters where GMD was higher in the CLBP group compared to HC (**Figure 4**). Similar results were obtained when using different smoothing kernels (sigma = 2 mm and sigma = 3 mm; see **Supplementary 3 and Supplementary 4)**.

**Figure 4:**
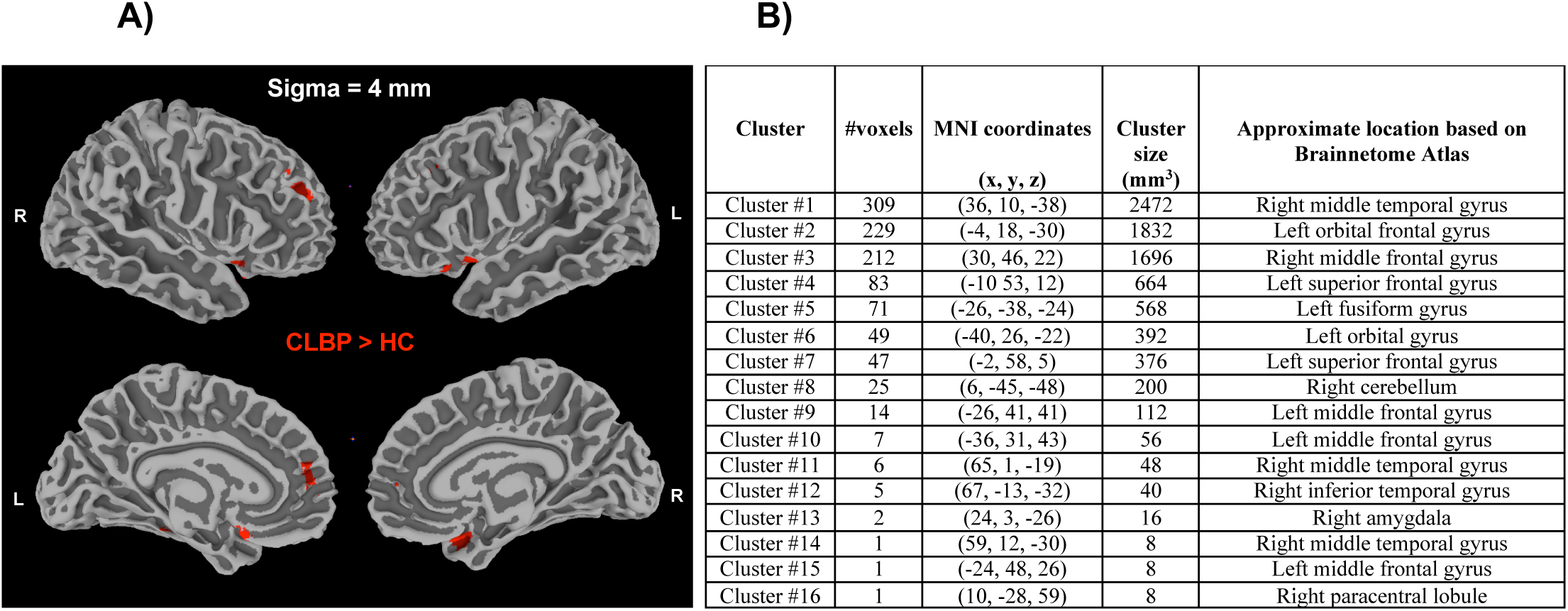
CLBP patients exhibit higher GMD in 16 clusters when intersecting voxels present across all three visits. Visual representation of the clusters that intersected across visits (**A**). A detailed description of the 16 clusters coordinates, size and label based on the Brainnetome Atlas (**B**).

Subsequently, after testing three different clustering (15, 30, and 45 voxels), the total number of clusters decreased by 50% (16 to 8), 56% (16 to 7), and 56 % (16 to 7), respectively. Cluster made of at least 45 voxels were retained. Similar results were obtained when using different smoothing kernels (sigma = 2 mm and sigma = 3 mm; see **Supplementary 3 and Supplementary 4)**.

Finally, after keeping only clusters present for each smoothing, three clusters were still present (**Figure 5**). Two clusters were in the frontal lobe (right middle frontal gyrus (r-MFG) and left orbital frontal gyrus (l-OFG)) and one in the medial temporal lobe (right middle temporal gyrus (r-MTG). A detailed description of these three clusters is provided in **Table 2, Supplementary 5, and Supplementary 6**. These findings demonstrate that after applying a stringent approach, GMD is higher in CLBP compared to HC.

**Figure 5:**
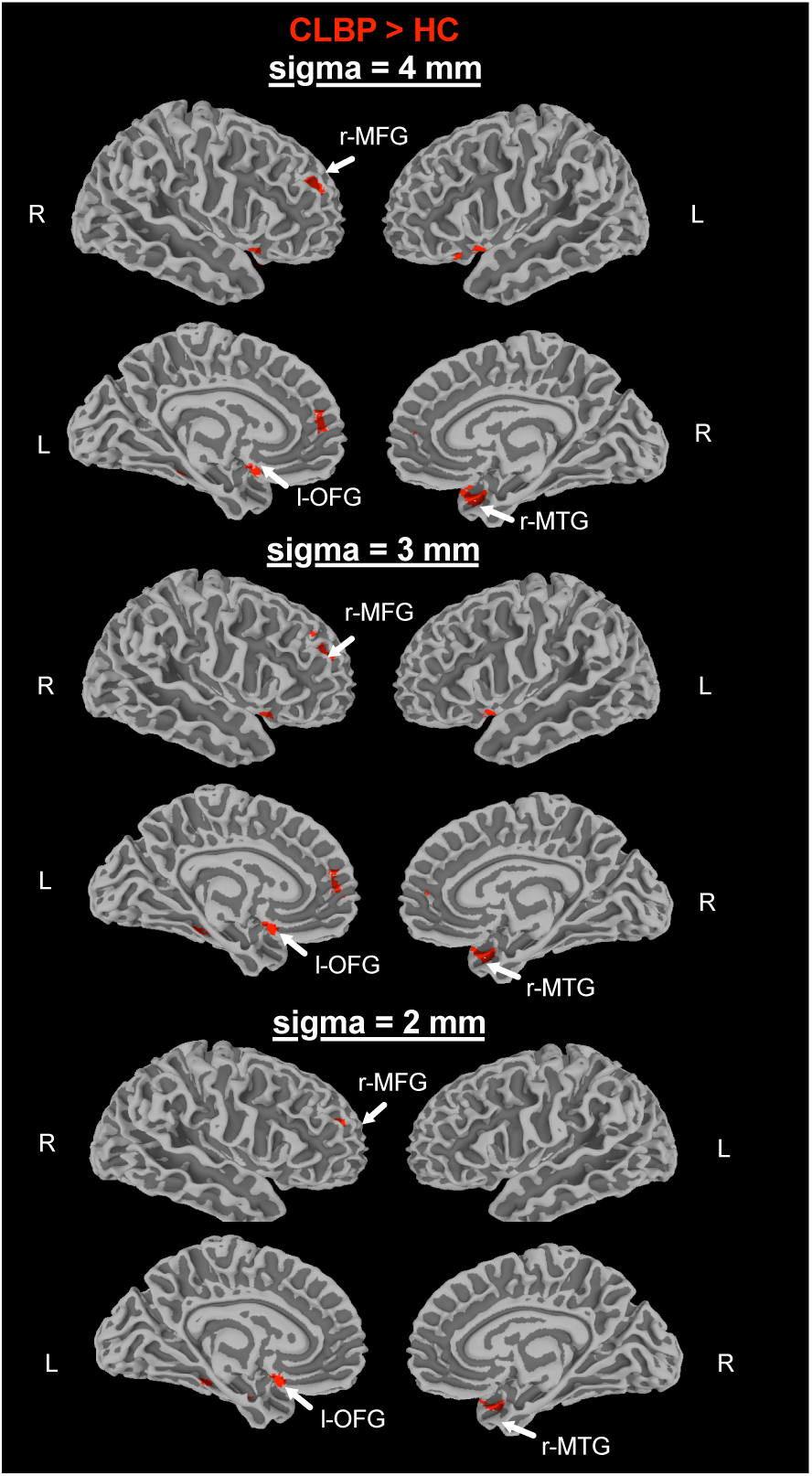
CLBP patients exhibit higher GMD in 3 clusters when keeping only clusters present at each smoothing. White arrows identified cluster present for each smoothing, which are found in the right middle temporal gyrus (r-MTG), left orbital frontal gyrus (l-OFG) and right middle frontal gyrus (r-MFG).

**Table 2:** A detailed description of the four remaining clusters (sigma = 4 mm)

| Cluster | # voxels | MNI coordinates (x, y, z) | Cluster size (mm<sup>3</sup>) | GMD (0-1) |  |  |  |  |  |
| --- | --- | --- | --- | --- | --- | --- | --- | --- | --- |
|  |  |  |  | BL |  | 2M |  | 4M |  |
|  |  |  |  | CLBP | HC | CLBP | HC | CLBP | HC |
| Cluster r-MTG | 309 | (36, 10, -38) | 2472 | 0.51±0.06 | 0.46±0.06 | 0.51±0.07 | 0.45±0.06 | 0.51±0.07 | 0.45±0.06 |
| Cluster l-OFG | 229 | (-4, 18, -30) | 1832 | 0.59±0.05 | 0.53±0.06 | 0.59±0.06 | 0.53±0.05 | 0.59±0.06 | 0.54±0.05 |
| Cluster r-MFG | 212 | (30, 46, 22) | 1696 | 0.50±0.06 | 0.45±0.06 | 0.51±0.05 | 0.45±0.06 | 0.52±0.06 | 0.45±0.06 |

r-MTG: right middle temporal gyrus
l-OFG: left orbital frontal gyrus
r-MFG: right middle frontal gyrus

### 3.4 CLBP patients do not show stable and reproducible regions with lower GMD

After applying the same methodological approach to evaluate the presence of significant regions with lower GMD in CLBP compared to HC, no voxels survived the full methodology (see **Supplementary 7, Supplementary 8, and Supplementary 9** for individual smoothing maps and cluster tables).

## 4. Discussion

In this study, we evaluated whether individuals with mild to moderate CLBP, who were not receiving prescribed pharmacological treatment, exhibit differences in their GMD compared to HC. We observed that individuals with CLBP had higher GMD than HC, particularly in the frontal and medial temporal lobes. Taken together, these results suggest that the observed GMD alterations may be related to the CLBP condition itself, and can be detected even in individuals with milder symptoms and higher physical function, without the confounding effects of opioids, anticonvulsants, or antidepressants.

This contrasts with the majority of previous studies, which have reported lower GMD in CLBP patients (Apkarian et al., 2004; Baliki et al., 2011; Ivo et al., 2013; Ung et al., 2014; Yu et al., 2021) – often interpreted as cortical atrophy (Apkarian et al., 2004; Marcori & Okazaki, 2019; Wang et al., 2017). However, it is important to reiterate that our sample differed from the literature in two ways: 1) participants had mild to moderate symptom severity (average pain intensity of 4/10, low pain disability based on similar questionnaire), compared to literature which is based on patients with moderate to high symptom severity (average pain intensity of 7/10 and moderate pain disability based on a pain disability questionnaire); and 2) participant had at no point taken pain medication such as opioids, anticonvulsants or antidepressants which are known to influence GM (Sean et al., 2024). Medications can affect cerebral gray matter volume as showed in Younger et al. (2012), wherein long-lasting reduced grey matter volume was observed in several brain regions following only one month of morphine treatment (Younger et al., 2011).

Together, this suggest that our results could be different from the literature because our CLBP patients likely present a different clinical profile (mild to moderate symptoms), and could be in a distinct brain state. While it is difficult to determine to what extent the difference in result is attributable to the difference in medication vs. the difference in symptom severity. For example, Maleki et al. (2013) showing that an increase hippocampus GMV in people with a low frequency of migraine attacks (i.e., a less severe form of the condition) compared to people with a high frequency of migraine attacks (32). These observations support the idea that the higher GMD we observed could be attributable – at least partly – to the lower symptom severity of our sample.

In terms of regional specificity, our most significant clusters of increased GMD were localized to the frontal lobe and in the medial temporal lobe. Other studies have reported abnormalities in regions similar to those observed in our CLBP patients, such as the frontal and medial temporal lobes (Apkarian et al., 2004; Baliki et al., 2011; Ivo et al., 2013). These regions appear to be crucial in the dynamic progression from the acute phase to the chronic phase, as well as in its maintenance (Ayoub et al., 2019; Baliki & Apkarian, 2015; McCarberg & Peppin, 2019; Ong et al., 2019; Stegemann et al., 2023; Thompson & Neugebauer, 2019; Waisman & Katz, 2024). The frontal lobe and medial temporal lobe play an important role in learning and memory (Jeneson & Squire, 2012; McDonald et al., 2001). Several studies suggest that CP could be a persistence of the memory of pain and/or the inability to extinguish the memory of pain evoked by an initial inciting injury (Apkarian, 2008; Apkarian et al., 2009; Biggs et al., 2022; Mansour et al., 2014; Phelps et al., 2021; Simons et al., 2014; Vachon-Presseau, 2018; Yi & Zhang, 2011). Stegemann et al. (2023) has highlighted the importance of frontal lobe in CP, by showing that a network of prefrontal cells encoding fear memories are involved in the development and maintenance of chronic pain states (Stegemann et al., 2023). Furthermore, other studies have shown that people who develop exaggerated memories of their past pain will tend to develop a memory bias towards their future pain and are more likely to develop CP (Baliki & Apkarian, 2015; Berger et al., 2018; Houde et al., 2020; Jeneson & Squire, 2012; McDonald et al., 2001; Salovey et al., 1993; Waisman et al., 2022; Waisman & Katz, 2024; Williams et al., 2022). Another study from Gui et al. (2016) has showed an increased expression of interleukin-1beta (IL-1β), an inflammatory marker, in several regions associated with pain, memory, and emotion, like the prefrontal cortex and medial temporal lobe (Gui et al., 2016).

It is important to mention that the biological interpretation of “GMD” remains uncertain in the literature. Eriksson et al. (2012) found no correlation between quantitative grey matter histological measures (i.e., neuronal density and glial fibrillary acidic protein) and grey matter probability maps (i.e., GMV/GMD). In contrast, Keifer et al. (2016) identified a correlation between dendritic spine density (which could reflect synapses) and grey matter probability maps. Another possible interpretation could involve neuroinflammation (Ransohoff, 2016), a phenomenon shown in some CLBP studies using Positron Emission Tomography (PET) imaging (Alshelh et al., 2022; Loggia, 2024; Loggia et al., 2015; Nutma et al., 2023). For instance, Loggia et al. (2015) and Torrado-Carvajal et al. (2021) have showed elevated brain levels of translocator protein (TSPO) in people with CLBP compared to HC, which is consider a marker of glial activation or density of inflammatory cells (Loggia, 2024; Nutma et al., 2023). Interestingly, patients in the studies by Loggia et al. (2015) and Torrado-Carvajal et al. (2021) had an average pain intensity level of 3/10, which is considered mild pain and similar to our patients (Schwerin & Mohney, 2024). Alternately, the increase in GMD could also reflect gliosis, a nonspecific reactive change in glial cells in response to damage to the central nervous system (CNS) (Burda & Sofroniew, 2014). In most cases, gliosis involves the proliferation or hypertrophy of several different types of glial cells, including astrocytes, microglia, and oligodendrocytes (Ko, 2020). Interestingly, a recent review from Emvalomenos et al. (2024) has mentioned that gliosis could be a promising avenue to study in the transition from acute to chronic pain (Emvalomenos et al., 2024). However, it is important to mention that on a T1- weighted image, it is difficult to conclude on a potential mechanism (see limitation). Additionally, there is a lack of post-mortem or histology studies on CP patients’ brains, which could have helped us clarify the underlying mechanism related to the observed difference in GMD.

### 4.1 Strengths and limitations

One of the strengths of our study is that we were able to examine GMD in CLBP patients with low to mild symptom severity, without the confounding influence of opioids, anticonvulsants or antidepressants. Another strength of our dataset lies in the longitudinal aspect, where each participant was imaged three times over a four-month period. We further took advantage of this repeated measure to develop a rigorous method to minimize false positive findings.

One of the limitations of our study is that we rely on T1-weighted images, which makes it difficult to interpret our results with certainty, specifically because the intensity in a T1-weighted image is not quantitative. Having other sequences such as a Fluid-Attenuated Inversion Recovery (FLAIR) or free-water derived from diffusion MRI, could have helped us to evaluate whether neuroinflammation was present or not in our CLBP patients.

Another limitation is the small sample size, which affects the statistical power and accordingly increases the likelihood of both of Type I and Type II errors. However, having three assessment time point allowed us to create a stringent methodological approach rarely seen in the MRI literature that was aimed at reducing both of Type I and Type II errors. Also, we only analyzed our results using one pipeline (FSL-VBM), but the software options allowing the construction of a template based on the anatomy of our participants are limited and Eggert et al. 2012 have shown that FSL achieved the highest accuracy compared to other software (Eggert et al., 2012). Another study from Johnson et al. 2017 have compared seven segmentation tools in HC and Huntington’s disease and shown that all methods showed high reliability (Johnson et al., 2017).

Finally, pain duration was not systematically collected for every patient (as a large portion of them were unable to give a specific answer), such that we were unable to properly account for pain duration in statistical analyses.

## 5. Conclusion

Our results suggest that individuals with CLBP, experiencing mild to moderate pain symptoms and not using centrally acting medications, exhibit increased GMD compared to HC, as assessed using a robust repeated measures approach. The most significant clusters were observed in the frontal lobe (two clusters) and the medial temporal lobe. However, since only T1-weighted images were acquired, the biological interpretation of these findings remains uncertain. Future studies should measure pain duration and ideally include patients with low, moderate, and high symptom severity, both with and without centrally acting medications, to better understand the contribution of each of these variables. Additionally, using other imaging modalities will help clarify the underlying mechanisms responsible for the observed differences in GMD.

## Data availability

Our data are available from the corresponding author upon request.

## Author Contributions

MS and PT conceived the study. MS, SC, KW and PT analyzed the results. MS, SC, ACL, JH, MM, GL, KW, PT contributed to writing the manuscript. MS, GL, KW, and PT helped with data analysis and interpretation. MS, MM and PT collected data.

## Data Availability

Our data are available from the corresponding author upon request.

