## Supplementary 1 for "Higher Brain Grey Matter Density in Mild to Moderate Chronic Low Back Pain Patients"

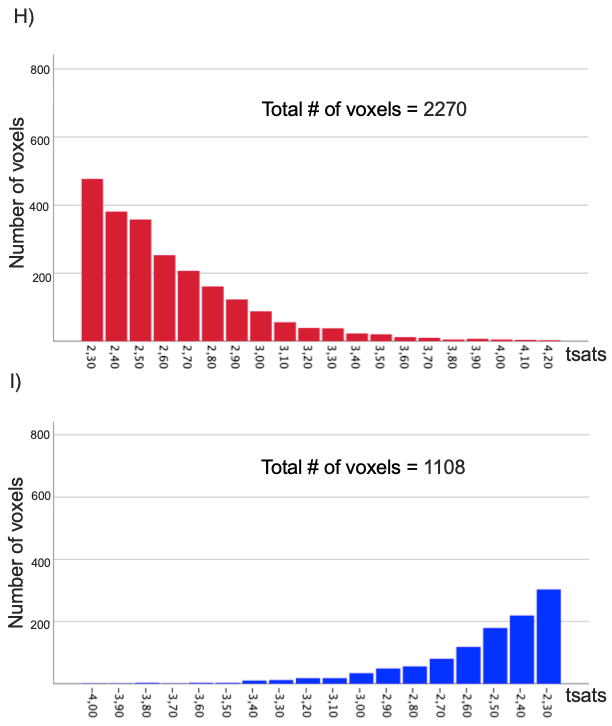

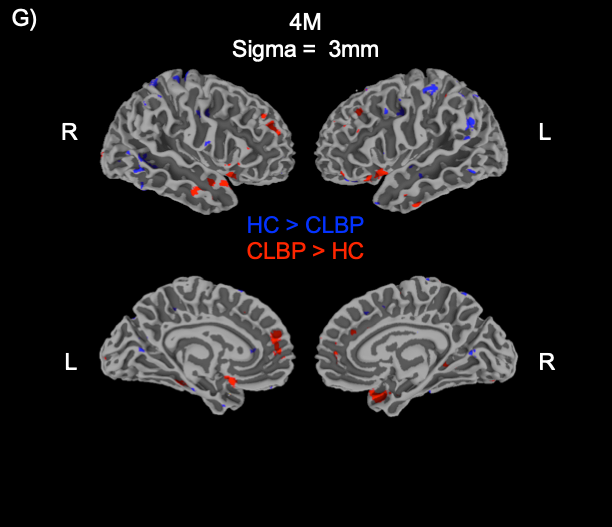

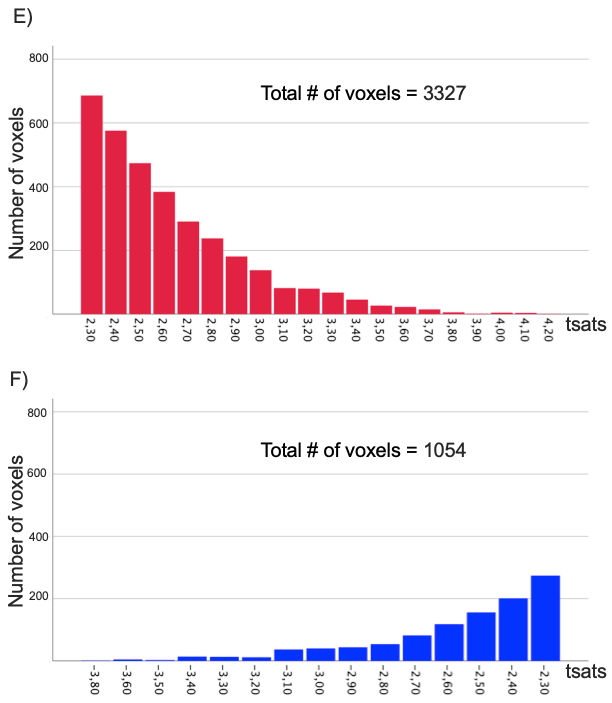

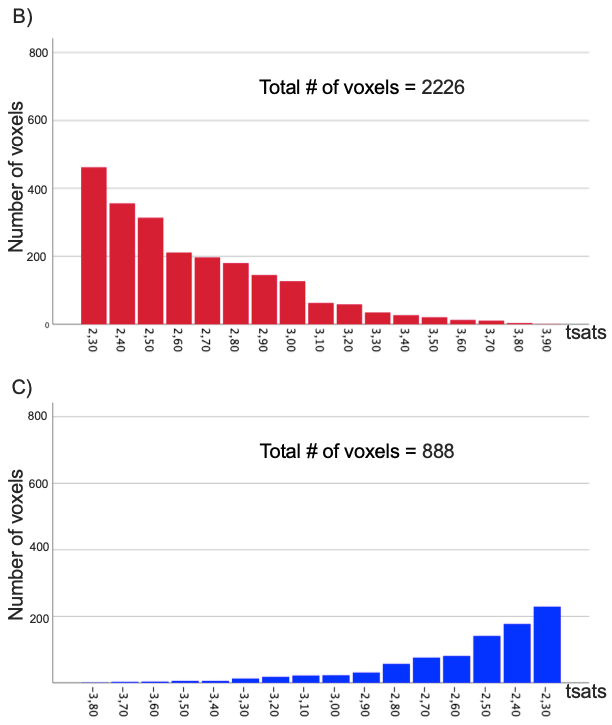

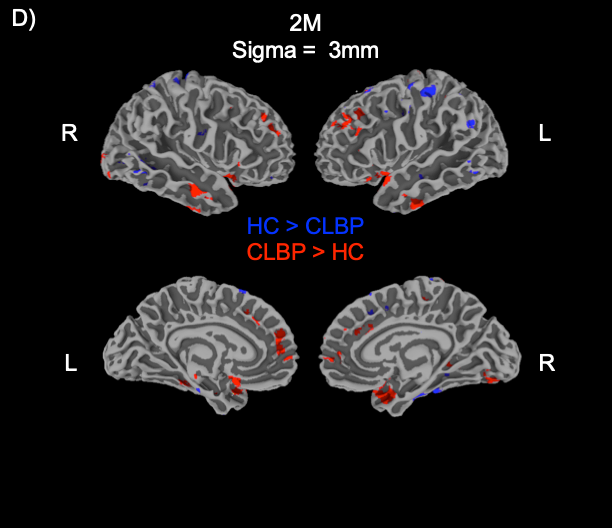

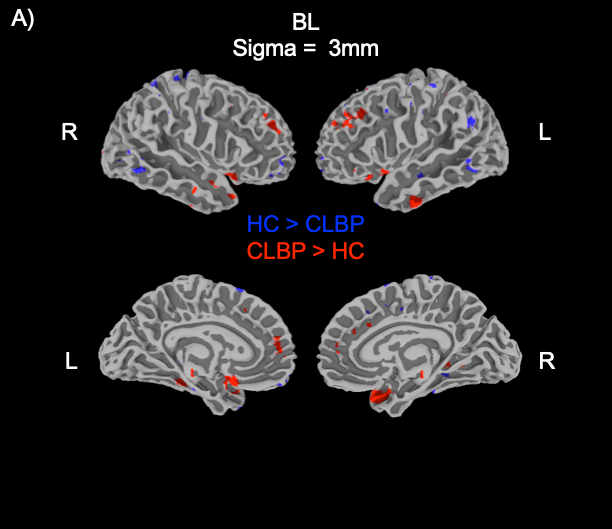

**Supplementary 1: CLBP patients show higher GMD (sigma = 3mm)**

GMD differences between groups are shown at A) BL, B) 2M and C) 4M. Voxels wherein GMD is lower in the CLBP group compared to HC are shown in blue. Voxels wherein GMD is higher in the CLBP group compared to HC are shown in red. For each map, two bar charts are presented to show the number of significant voxels for each t-statistics. The red bar chart (B, E, and H) indicates the number of voxels wherein GMD is higher in the CLBP group compared to HC (t ≥ 2.3) at BL, 2M, and 4 M respectively. The blue chart (C, F, and I) indicated the number of voxels wherein GMD is lower in the CLBP group compared to HC (t ≥ -2.3) at BL, 2M, and 4 M respectively.From all the statistically significant voxels, 71%, 76% and 67% showed higher GMD in CLBP compared to HC, respectively at BL, 2M and 4M.
