## Supplementary 3 for "Higher Brain Grey Matter Density in Mild to Moderate Chronic Low Back Pain Patients"

**A)	B)**

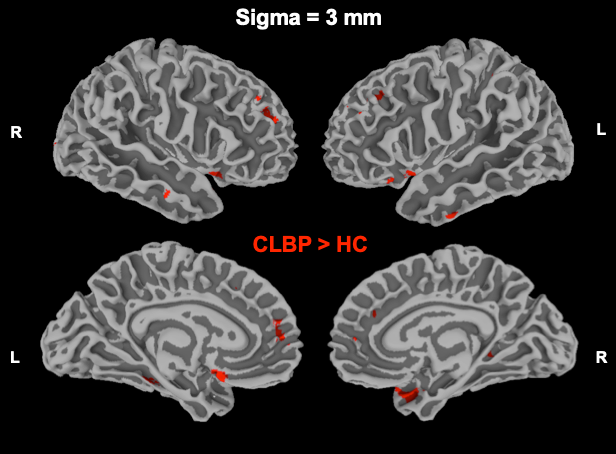

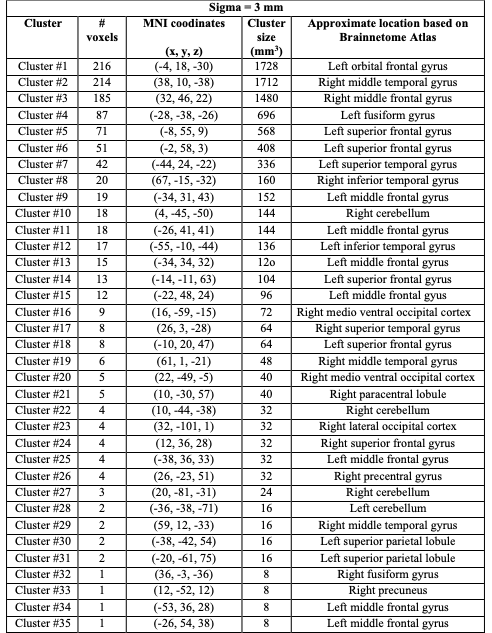

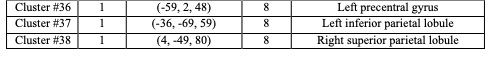

**Supplementary 3: CLBP patients exhibit higher GMD in 38 clusters when intersecting voxels present across all three visits (sigma = 3 mm).** Visual representation of the clusters that intersected across visits **(A**). A detailed description of the 38 clusters coordinates, size and label based on the Brainnetome Atlas (**B**). After testing three different clustering (15, 30, and 45 voxels), the total number of clusters was reduced from 66% (38 to 13), 82% reduction (38 to 7), and 84 % reduction (38 to 6), respectively.
