## Supplementary 4 for "Higher Brain Grey Matter Density in Mild to Moderate Chronic Low Back Pain Patients"

A) **B)**

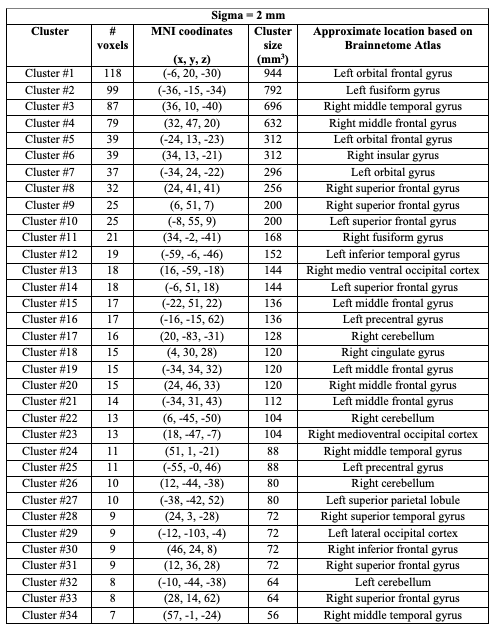

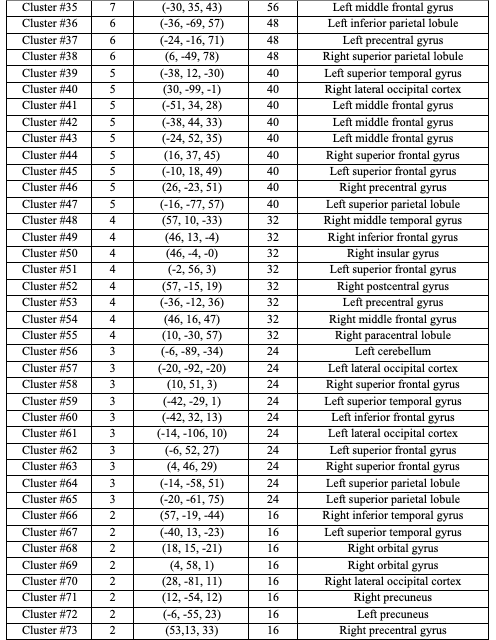

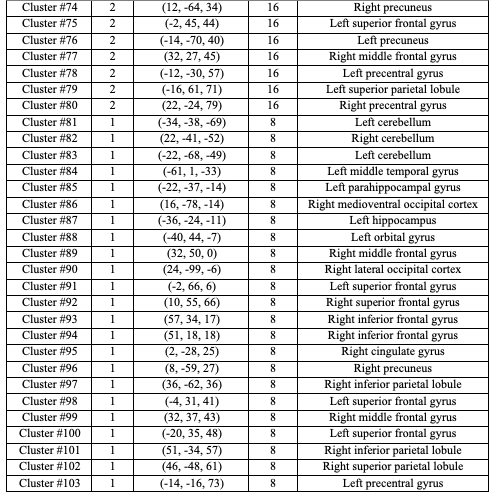

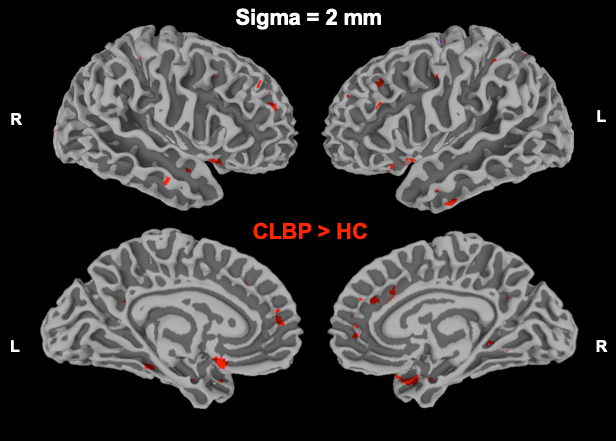

**Supplementary 4: CLBP patients exhibit higher GMD in 103 clusters when intersecting voxels present across all three visits (sigma = 2 mm).** Visual representation of the clusters that intersected across visits **(A**). A detailed description of the 103 clusters coordinates, size and label based on the Brainnetome Atlas (**B**). After testing three different clustering (15, 30, and 45 voxels), the total number of clusters was reduced from 81% (103 to 20), 92% reduction (103 to 8), and 96 % reduction (103 to 4), respectively.
