## Supplementary 5 for "Higher Brain Grey Matter Density in Mild to Moderate Chronic Low Back Pain Patients"

**Supplementary 5: A detailed description of the three remaining clusters (sigma = 3 mm)**

| **Cluster** | **# voxels** | **MNI coodinates (x, y, z)** | **Cluster size (mm^3^)** | **GMD (0-1)** | | | | | |
| --- | --- | --- | --- | --- | --- | --- | --- | --- | --- |
|  |  |  |  | **BL** | | **2M** | | **4M** | |
|  |  |  |  | **CLBP** | **HC** | **CLBP** | **HC** | **CLBP** | **HC** |
| Cluster  r-MTG | 214 | (38, 10, -38) | 1712 | 0.57±0.08 | 0.49±0.07 | 0.57±0.08 | 0.50±0.07 | 0.57±0.09 | 0.50±0.07 |
| Cluster l-OFG | 216 | (-4, 18, -30) | 1728 | 0.64±0.06 | 0.57±0.07 | 0.64±0.08 | 0.57±0.06 | 0.64±0.07 | 0.57±0.06 |
| Cluster  r-MFG | 185 | (32, 46, 22) | 1480 | 0.57±0.06 | 0.50±0.07 | 0.57±0.06 | 0.51±0.06 | 0.58±0.07 | 0.51±0.06 |

r-MTG: right middle temporal gyrus

l-OFG: left orbital frontal gyrus

r-MFG: right middle frontal gyrus
