## Supplementary 6 for "Higher Brain Grey Matter Density in Mild to Moderate Chronic Low Back Pain Patients"

**Supplementary 6: A detailed description of the three remaining clusters (sigma = 2 mm)**

| **Cluster** | **# voxels** | **MNI coodinates (x, y, z)** | **Cluster size (mm^3^)** | **GMD (0-1)** | | | | | |
| --- | --- | --- | --- | --- | --- | --- | --- | --- | --- |
|  |  |  |  | **BL** | | **2M** | | **4M** | |
|  |  |  |  | **CLBP** | **HC** | **CLBP** | **HC** | **CLBP** | **HC** |
| Cluster  r-MTG | 87 | (36, 10, -40) | 696 | 0.65±0.10 | 0.54±0.09 | 0.66±0.10 | 0.55±0.09 | 0.65±0.11 | 0.55±0.09 |
| Cluster l-OFG | 118 | (-6, 20, -30) | 944 | 0.72±0.09 | 0.62±0.09 | 0.72±0.11 | 0.63±0.08 | 0.72±0.10 | 0.63±0.09 |
| Cluster  r-MFG | 79 | (32, 47, 20) | 632 | 0.61±0.08 | 0.52±0.09 | 0.61±0.08 | 0.52±0.08 | 0.63±0.09 | 0.52±0.07 |

r-MTG: right middle temporal gyrus

l-OFG: left orbital frontal gyrus

r-MFG: right middle frontal gyrus
