## Supplementary 7 for "Higher Brain Grey Matter Density in Mild to Moderate Chronic Low Back Pain Patients"

**A)	B)**

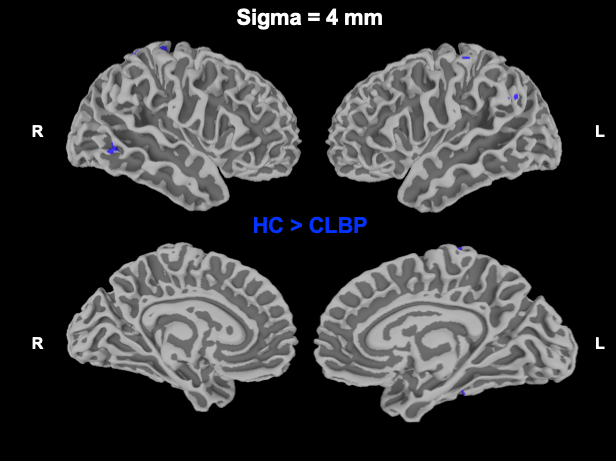

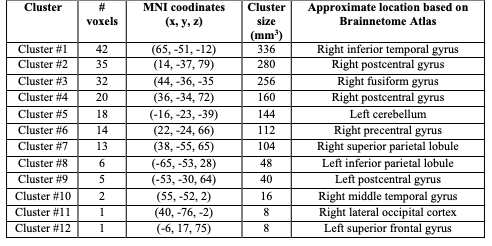

**Supplementary 7:** **CLBP patients exhibit lower GMD in 12 clusters when intersecting voxels present across all three visits.** Visual representation of the clusters that intersected across visits (**A**). A detailed description of the 12 clusters coordinates, size and label based on the Brainnetome Atlas (**B**). After testing three different clustering (15, 30, and 45 voxels), the total number of clusters was reduced from 58% (12 to 5), 75% reduction (12 to 3), and 100 % reduction (12 to 0), respectively.
