## Supplementary 8 for "Higher Brain Grey Matter Density in Mild to Moderate Chronic Low Back Pain Patients"

**A)	B)**

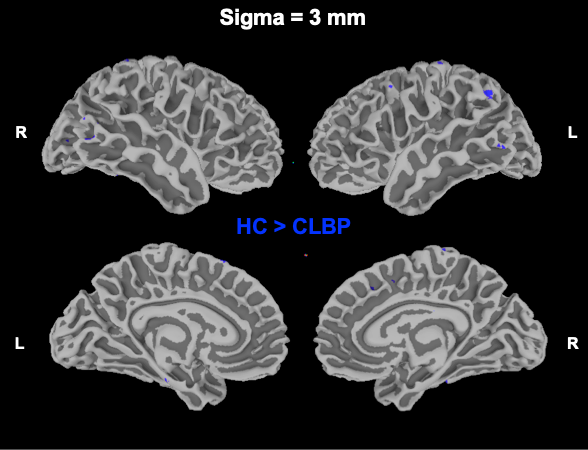

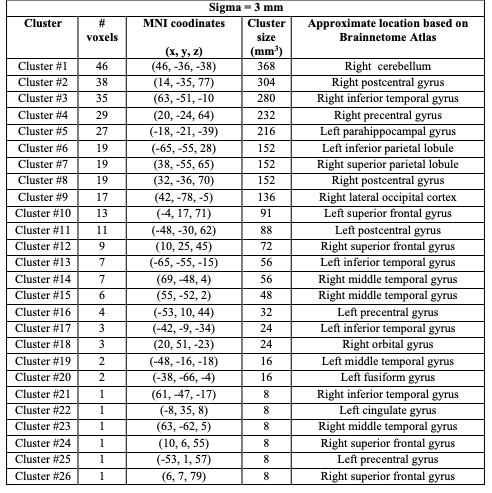

**Supplementary 8: CLBP patients exhibit lower GMD in 26 clusters when intersecting voxels present across all three visits (sigma = 3 mm).** Visual representation of the clusters that intersected across visits **(A**). A detailed description of the 26 clusters coordinates, size and label based on the Brainnetome Atlas (**B**). After testing three different clustering (15, 30, and 45 voxels), the total number of clusters was reduced from 65% (26 to 9), 88% reduction (26 to 3), and 96% reduction (26 to 1), respectively.
