## Supplementary 9 for "Higher Brain Grey Matter Density in Mild to Moderate Chronic Low Back Pain Patients"

**A)	B)**

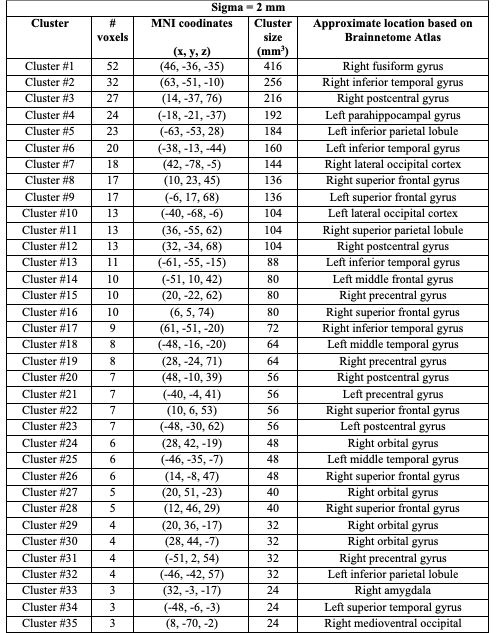

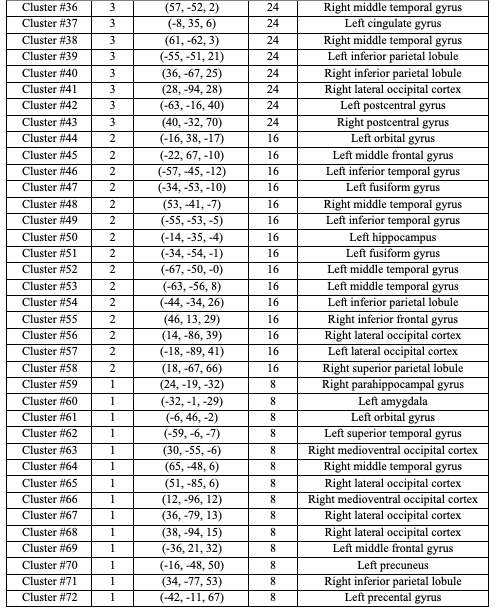

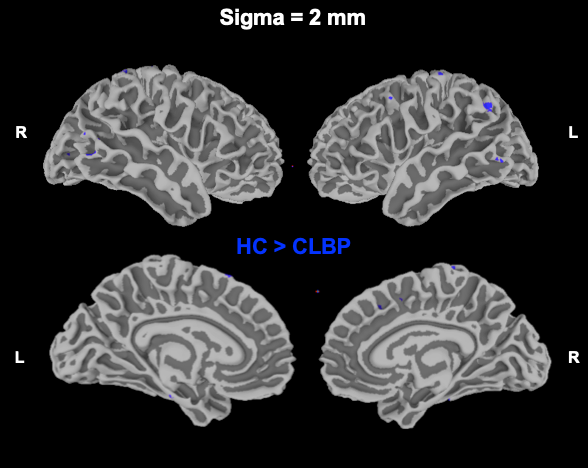

**Supplementary 9: CLBP patients exhibit lower GMD in 72 clusters when intersecting voxels present across all three visits (sigma = 3 mm).** Visual representation of the clusters that intersected across visits **(A**). A detailed description of the 72 clusters coordinates, size and label based on the Brainnetome Atlas (**B**). After testing three different clustering (15, 30, and 45 voxels), the total number of clusters was reduced from 88% (72 to 9), 99% reduction (72 to 1), and 99 % reduction (72 to 1), respectively.
